# Estimating the risk of incident SARS-CoV-2 infection among healthcare workers in quarantine hospitals: the Egyptian example

**DOI:** 10.1101/2020.12.21.20248594

**Authors:** Sofía Jijón, Ahmad Al Shafie, Essam Hassan, EMAE-MESuRS working group on nosocomial SARS-CoV-2 modeling, Laura Temime, Kévin Jean, Mohamed El-Kassas

## Abstract

In response to the COVID-19 epidemic, Egypt established a unique care model based on quarantine hospitals where only externally-referred confirmed COVID-19 patients were admitted, and healthcare workers resided continuously over 1-to 2-week working shifts. While the COVID-19 risk for HCWs has been widely reported in standard healthcare settings, it has not been evaluated yet in quarantine hospitals.

Here, we relied on longitudinal data, including results of routine RT-PCR tests, collected within three quarantine hospitals located in Cairo and Fayoum, Egypt. Using a model-based approach that accounts for the time-since-exposure variation in false-negative rates of RT-PCR tests, we computed the incidence of SARS-CoV-2 infection among HCWs. Over a total follow-up of 6,064 person-days (PD), we estimated an incidence rate (per 100 PD) of 1.05 (95% CrI: 0.58–1.65) at Hospital 1, 1.92 (95% CrI: 0.93–3.28) at Hospital 2 and 7.62 (95% CrI: 3.47–13.70) at Hospital 3. The probability for an HCW to be infected at the end of a shift was 13.7% (95% CrI: 7.8%–20.8%) and 23.8% (95% CrI: 12.2%–37.3%) for a 2-week shift at Hospital 1 and Hospital 2, respectively, which lies within the range of risk levels previously documented in standard healthcare settings, whereas it was >3-fold higher for a 7-day shift at Hospital 2 (42.6%, 95%CrI: 21.9%–64.4%). Our model-based estimates unveil a proportion of undiagnosed infections among HCWs of 46.4% (95% CrI: 18.8%–66.7%), 45.0% (95% CrI: 5.6%–70.8%) and 59.2% (95% CrI: 34.8%–78.8%), for Hospitals 1 to 3, respectively.

The large variation in SARS-CoV-2 incidence we document here suggests that HCWs from quarantine hospitals may face a high occupational risk of infection, but that, with sufficient anticipation and infection control measures, this risk can be brought down to levels similar to those observed in standard healthcare settings.

**WHAT THIS PAPER ADDS:** *What is already known on this topic:* Previous studies conducted in standard care settings have documented that frontline healthcare workers (HCWs) face high risk of COVID-19. Whether risk levels differ in alternative care models, such as COVID-19 quarantine hospitals in Egypt where HCWs resided in the hospital days and nights for various durations, is unknown.

*What this study adds:* COVID-19 risk for HCWs in quarantine hospitals varies substantially between facilities, from risk levels that are in the range of those documented in standard healthcare settings to levels that were approximatively 3 times higher.

*How this study might affect research, practice or policy:* With sufficient anticipation and infection control measures, occupational COVID-19 risk for HCWs working in quarantine hospitals can be brought down to levels similar to those observed in standard healthcare settings.

## INTRODUCTION

Healthcare settings have faced multiple challenges related to the currently ongoing severe acute respiratory syndrome coronavirus 2 (SARS-CoV-2) pandemic. They have notably had to deal with influxes of infected patients across successive epidemic waves while controlling the risk of nosocomial spread to other patients and staff. As a result, healthcare workers (HCWs) have been a population of interest in terms of risk assessment and control measure implementation [1].

At early stages of the pandemic, Egypt was identified as one of the African countries most vulnerable to SARS-CoV-2 importation [2]. On February 14, 2020, Egypt reported the first confirmed case of coronavirus disease 2019 (COVID-19) in Africa and remained among the five African countries most assigned by the COVID-19 epidemic up to the end of 2020 [3]. From February 14 to August 31, 2020 (the period to which we refer to as the first wave), the COVID-19 epidemic in Egypt resulted in about 9,700 confirmed infections and 5,500 deaths reported nationally [4]. These numbers most certainly reflect underreporting of the real number of infections, due to the high proportion of asymptomatic SARS-CoV-2 infection [5,6] and limited testing.

To mitigate the high risk of SARS-CoV-2 spreading, Egypt established a unique care model under supervision from the World Health Organization, whereby specific hospitals were assigned as quarantine hospitals for patients with COVID-19, and where dedicated medical teams stayed in the hospital days and nights during working shifts of various durations [7].

While the quarantine-hospital strategy has the potential to be highly efficient in terms of patient care, as well as in limiting the potential spread of the virus from hospitals into the community, its impact on infection risk for HCWs remains understudied. One theoretical modelling study assessing healthcare working force organization found that alternating HCW teams by 1-week periods may reduce the overall number of infected HCWs [8]. On the other hand, to the best of our knowledge, previous epidemiological studies focusing on the infection risk faced by Egyptian HCWs have been conducted in non-quarantine settings exclusively [9– 11]. Yet, the quarantine strategy was adopted again to face the second wave of the COVID-19 epidemic at the national level, and could be adopted for future epidemic waves in Egypt and/or in other countries [12].

Here, we used mathematical modelling to estimate the risk of SARS-CoV-2 infection among HCWs participating in quarantine-hospital interventions, relying on detailed longitudinal data collected in three Egyptian healthcare facilities during the first wave of the COVID-19 epidemic.

## METHODS

### Study settings

Data was collected within three Egyptian hospitals (hereafter denoted by Hosp1, Hosp2 and Hosp3) located in Cairo (Hosp1 and Hosp3), and Fayoum (middle Egypt, Hosp2), that were temporarily transformed into quarantine hospitals during the first COVID-19 wave. During the quarantine-organization period (Hosp1: March 14th to August 1st, 2020; Hosp2: April 1st to July 31th; Hosp3: June 6th to July 11th, 2020), only externally-referred COVID-19 confirmed patients were admitted to these hospitals for medical care. Multidisciplinary medical teams fully dedicated to COVID-19 patient management worked in total isolation, organized by shifts (Hosp1: 2-week shifts; Hosp2: 1 to 2-week shifts; Hosp3: 1-week shifts). During their shift, HCWs were assigned either to intensive care units (ICU) or non-ICU. HCWs were screened for SARS-CoV-2 infection before starting a working shift using rapid serological IgM/IgG antibody tests (Artron laboratories Burnaby, Canada; sensitivity: 83.3%, specificity: 100% [13]). Only HCWs with no SARS-CoV-2 antibodies were allowed to start working in the hospital; except for the last shift in Hosp3, where staff recruitment relied exclusively on HCWs who had been previously infected (positive serological tests), and were additionally tested using reverse transcription-polymerase chain reaction (RT-PCR) on nasopharyngeal swabs, obtaining negative results. During the working shifts, HCWs were tested for SARS-CoV-2 infection using RT-PCR tests: i) routinely at the end of the shift, ii) upon symptoms, and iii) in case of outbreak suspicion (>2 positive tests among HCWs). HCWs testing negative at the end of the shift were then released for self-isolation at home for two weeks. HCWs testing positive before or during their shift self-isolated at home in the case of presenting no or mild symptoms, or were admitted to the same quarantine hospital for medical care in the case of presenting moderate to severe symptoms.

### Observed risk of SARS-CoV-2 infection

Crude incidence rates were obtained from the number of incident SARS-CoV-2 infections observed (ie, diagnosed during or at the end of a shift) among HCWs screened as seronegative for SARS-CoV-2 before their working shift. Incidence rate ratios were obtained through a Poisson regression adjusted by hospital and by type of care unit (ICU/ non-ICU). In addition, we computed the attack rates over each working shift, based on the observed infections among HCWs having a negative screening test at the beginning of the working shift. Observed per-shift attack rates of were considered to be healthcare-associated outbreaks among HCWs.

### Model-based estimates of the incidence rate of SARS-CoV-2 infection

An important feature arising from COVID-19 surveillance within quarantine hospitals is potential right-truncation of data: HCWs infected a short time before the end of a shift are likely to remain undiagnosed despite the systematic testing at the end of shifts, especially if the test is performed early in the incubation period. We thus developed the following mathematical model to estimate the incidence rate of both diagnosed and undiagnosed SARS-CoV-2 infections.

We simulated the daily number of incident infections among HCWs, 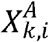 based on an unobserved binomial process:

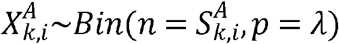

where *k* denotes the working shift, *i* denotes the day within the shift, *A* is the hospital unit of HCWs ’ assignments (*A* ∈ {ICU Non ICU}), 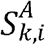 denotes the number of susceptible HCWs assigned to A, at the beginning of the i-th day of shift *k* and, *λ* denotes the constant daily probability of SARS-CoV-2 infection faced by HCWs. The number of susceptible HCWs was updated on a daily basis:

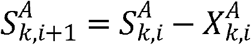

To model right-truncation in the surveillance data, we simulated an observation process representing the systematic RT-PCR testing of HCWs at the end of each shift, accounting for documented variation in test sensitivity as a function of time since infection [14]. The period of time since infection was computed as *j*_*k*_ *-i*, where *j*_*k*_ denotes the day at the end of shift k (i.e., the day when testing was performed) and i denotes the day of infection. Then, the daily number of incident infections eventually diagnosed at the end of the -th shift was given by

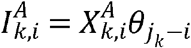

where 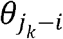 denotes the probability of testing positive to RT-PCR, *j* _*k*_*-i* days after infection.

We used a Bayesian Markov Chain Monte-Carlo (MCMC) approach to estimate the parameter λ that best fitted the observed number of infections. We assumed that λ was constant over the study period. We considered a non-informative uniform prior distribution ∼*U*(0,1) for λ and the following binomial likelihood function:

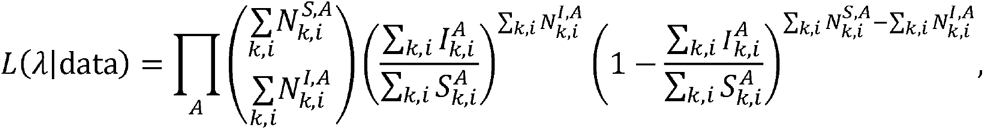

where 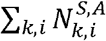 and 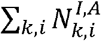 denote the total number of susceptible HCWs (denoted by the superscript *S*) and the total number of HCW infected and diagnosed with COVID-19 (denoted by the superscript *I*), observed in hospital unit *A*, during the study period, respectively. The model was thus fitted to the total number of observed infections in each hospital unit.

Medians and 95% credibility intervals (CrI) for λ were computed from posterior samples obtained after 10,000 model runs. The chains were visually inspected for convergence. For each hospital, we further estimated the probability for a susceptible HCW to be infected at the end of a shift as

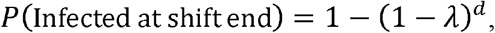

where *d* is the length of the working shift, in days.

Finally, to assess the proportion of undetected infections among HCWs, we ran our model on each hospital, using the posterior distributions for λ, to obtain an estimate for the real number of SARS-CoV-2 infections in each hospital (both diagnosed and undiagnosed), over the study period.

The numeric implementation of the model was coded in R 4.0.3 [15], using the FME package [16] for parameter estimation.

### Ethical approval

This study was approved by the Research Ethics Committee (REC) of the Central Directorate of Research and Health Development and Reviews at the Egyptian Ministry of Health and Population (Serial: 25-2020/16), and by REC for human subject research at the Faculty of Medicine, Helwan University (Serial: 50-2020).

## RESULTS

The study period covered ten 2-week shifts in Hosp1, nine ∼2-week shifts in Hosp2 and five 1-week shifts in Hosp3. The mean (min–max) number of HCWs per shift was 46 (34–63) in Hosp1, 15 (5–26) in Hosp2 and 19 (16–20) in Hosp3 (Table 1). Over a total follow-up of 8,733 person-days (PD), 54 SARS-CoV-2 infections were observed (ie., diagnosed by RT-PCR) among 722 HCWs showing no evidence of SARS-CoV-2 antibodies at the beginning of their shifts, across the three hospitals. This represented an overall incidence rate of 0.62 (95% CI: 0.45–0.78) diagnosed SARS-Cov-2 infections per 100 PD (Table 2). A significantly higher incidence rate was observed for HCWs working at Hosp3 as compared to the other two hospitals; and HCWs working in non-ICU units tended to be more at risk than HCWs working in ICU, though this difference was not statistically significant (Table 2).

**Table 1.** Hospital characteristics. Quarantine organization period, location and mean number of HCWs and patients by hospital (Hosp1, Hosp2 and Hosp3). Abbreviations: HCWs = Healthcare workers, PD = person-days.

|  | Hosp1 | Hosp2 | Hosp3 |
| --- | --- | --- | --- |
| **Quarantine organization period** | March 14 – August 1 | April 1 – July 31 | June 6 – July 11 |
| **Location** | Cairo | Fayoum | Cairo |
| **Mean daily number of patients (min–max)** | 62 (0–108) | 37 (0–103) | 8 (0–20) |
| **Mean number of HCWs per shift (min–max)** | 46 (34–63) | 15 (5–26) | 19 (16–20) |
| **Shift duration** | 14 days | 7–14 days | 7 days |

**Table 2.** Observed risk of SARS-CoV-2 infection among HCWs, by hospital and by hospital unit. A total of 54 infections were observed (ie., diagnosed before or at the end of a shift) over 8,733 person-days, in the three hospitals (Hosp1, Hosp1 and Hosp3). Crude rates of SARS-CoV-2 infection among HCWs by hospital and by hospital unit: intensive and non-intensive care units and 95% confidence intervals. A Poisson regression adjusted by hospital and by hospital unit was performed on the observed infections to obtain the incidence rate ratios and 95% confidence intervals. Abbreviations: CI = confidence interval, ICU = intensive care unit, IRR = incidence rate ratio, PD = person-days.

|  |  | Observations |  | Crude rates |  | Adjusted Poisson Regression |  |  |
| --- | --- | --- | --- | --- | --- | --- | --- | --- |
|  |  | Events | PD | Rate per 100 PD | 95% CI | IRR | 95% CI | *p* |
| **Hospital** | **Hosp1** | 28 | 6258 | 0.45 | 0.28–0.61 | 1 | ref | – |
|  | **Hosp2** | 11 | 1808 | 0.61 | 0.25–0.97 | 1.49 | 0.74–2.87 | 0.28 |
|  | **Hosp3** | 15 | 667 | 2.25 | 1.1–3.39 | 5.59 | 3.13–10.01 | <0.001 |
| **Hospital Unit** | **ICU** | 15 | 2628 | 0.57 | 0.28–0.86 | 1 | ref | – |
|  | **Non-ICU** | 39 | 6105 | 0.64 | 0.44–0.84 | 1.49 | 0.85–2.59 | 0.16 |

Healthcare-associated outbreaks among HCWs (observed attack rates of) represented ∼70% (38/54) of all SARS-CoV-2 infections observed over the study period (Figure 1). Two of these outbreaks occurred in Hosp2 with 30% (3/10) and 24% (5/21) of HCWs being infected over two different working shifts. An outbreak where 36% of HCWs were infected (16/44) occurred in Hosp1 around the same time period. Two outbreaks took place later on in Hosp3, leading to, respectively, 30% (6/20) and 40% (8/20) of susceptible HCWs being infected over two different shifts. Of note, each outbreak resulted in infections in both ICU and non-ICU. For the last shift in Hosp3, staff recruitment relied exclusively on HCWs who had been infected during the first or the second shifts. This likely prevented HCWs in Hosp3 from becoming infected during the last shift (cf. Figure 1).

**Figure 1.**
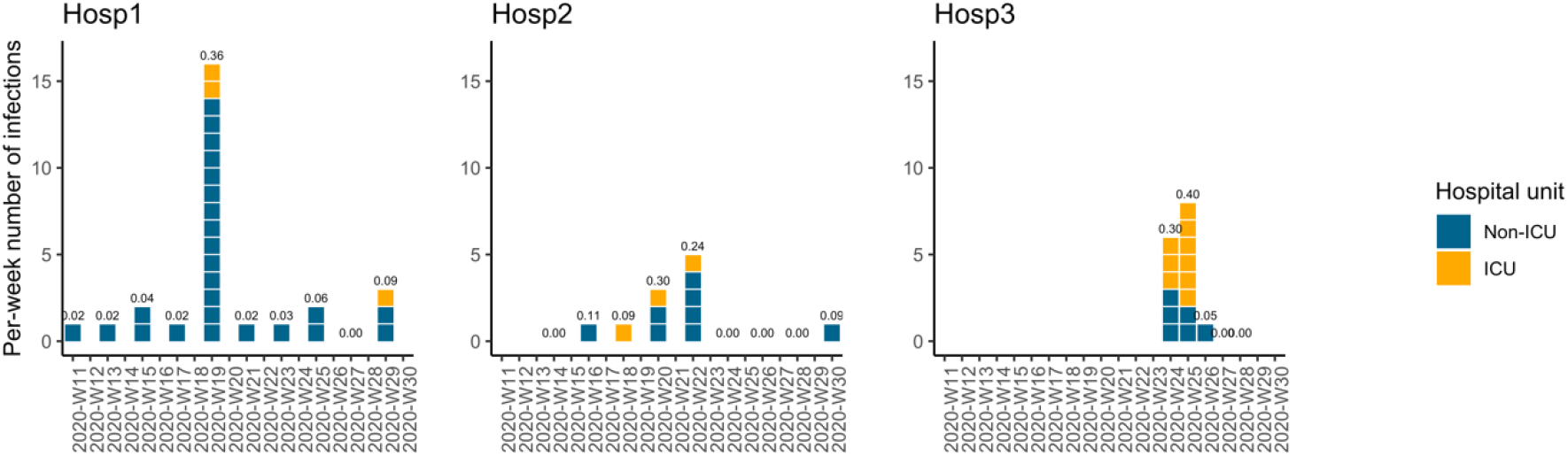
Per-week number of infections by hospital unit. For a given working shift, healthcare workers were assigned to either the intensive care unit (ICU, yellow) or a non-ICU (blue). Dates are given in weeks, denoted by the prefix W followed by the week number of year 2020. Hosp1 was established as a quarantine hospital from March 14th to August 1st (W11 to W31), Hosp2 from April 1st to July 31th (W14 to W31) and Hosp3 from June 6th to July 11th (W23 to W28). In Hosp1 and Hosp2, most infections occurred in non-ICU units. Per-shift attack rates are shown above the bars. Of note, only previously infected HCWs worked during the last two shifts (W27 and W28) in Hosp3.

To estimate the mean incidence rate of both observed and unobserved infections in each hospital, we ran our model considering a constant risk,, over the whole study period, in each hospital unit. The chains for λ, for the three hospitals, are depicted in the Supplementary Figure 1. We estimated significantly different risk levels between the three hospitals (Kolmogorov-Smirnov test; p-value), with an incidence rate of 1.05 (95% CrI: 0.58–1.65) per 100 PD in Hosp1, 1.92 (95% CrI: 0.93–3.28) per 100 PD in Hosp2 and 7.62 (95% CrI: 3.47–13.70) per 100 PD in Hosp3.

The probability for a HCW to be infected at the end of a working shift was estimated at 13.7% (95% CrI: 7.8%–20.8%) and 23.8% (95% CrI: 12.2%–37.3%) for 2-week shifts at Hosp1 and Hosp2, respectively, whereas a much higher probability of 42.6% (95% CrI: 21.9%–64.4%) was found for a 7-day shift at Hosp3 (see Table 3). These probabilities become 7.1% (95% CrI: 4.0%–11.0%) and 12.7% (95% CrI: 6.3%–20.8%), for Hosp1 and Hosp2, respectively, if 7-day shifts are considered as well, for easier comparison (Table 3). Given the notable differences between hospitals (Figure 1), we did not assess infection risk by care unit across hospitals, but within each hospital.

**Table 3.** Model-based estimates of all SARS-CoV-2 infections. The model-based estimates of the incidence rate, the probability of infection for a 7-day working shift and the estimated number of SARS-CoV-2 infections in each hospital, over the study period. Running our model for each type of care unit (ICU and non-ICU) in each hospital unveils the difference in the risk faced by HCWs between the three hospitals of our study. Indeed, HCWs working in Hosp1 and Hosp2 face a higher risk when assigned to non-ICU, as compared with those working in the ICU; whereas HCWs in Hosp3 face a slightly higher risk when assigned to the ICU. All estimates concern both diagnosed and undiagnosed infections (all), unless otherwise stated (diagnosed). Abbreviations: CrI = credibility interval, ICU = intensive care unit, PD = person-days.

|  | Median estimated incidence rate<br>per 100 PD (95% CrI) |  |  | Probability of<br>infection for a 7-day shift (95%<br>CrI) | Median number of<br>SARS-CoV-2 infections<br>(95% CrI) |  |  |
| --- | --- | --- | --- | --- | --- | --- | --- |
|  | ICU | Non-ICU | Overall |  | All | Diagnosed | Proportion of<br>undiagnosed<br>infections |
| **Hosp1** | 0.59<br>(0.11–1.61) | 1.27<br>(0.68–2.01) | 1.05<br>(0.58–1.65) | 7.1%<br>(4.0%–11.0%) | 59<br>(45–74) | 31<br>(15–51) | 46.4%<br>(18.8%–66.7%) |
| **Hosp2** | 1.33<br>(0.33–3.33) | 2.75<br>(1.12–5.02) | 1.92<br>(0.93–3.28) | 12.7%<br>(6.3%–20.8%) | 31<br>(14–52) | 17<br>(7–31) | 45.0%<br>(5.6%–70.8%) |
| **Hosp3** | 9.37<br>(3.48–19.66) | 7.12<br>(2.06–18.71) | 7.62<br>(3.47–13.70) | 42.6%<br>(21.9%–64.4%) | 41<br>(19–63) | 16<br>(6–30) | 59.2%<br>(34.8%–78.8%) |

The posterior distributions for the incidence rate λ yield the following model-based median numbers of total SARS-CoV-2 infections: 59 (95% CrI: 45–74) at Hosp1, 31 (95% CrI: 14– 52) at Hosp2 and 41 (95% CrI: 19–63) at Hosp3 (Table 3), which unveils a proportion of undiagnosed infections among HCWs of 46.4% (95% CrI: 18.8%–66.7%), 45.0% (95% CrI: 5.6%–70.8%) and 59.2% (95% CrI: 34.8%–78.8%), respectively.

We further ran our model on each hospital unit, on each hospital, which confirmed a notable difference between Hosp1–2 and Hosp3: our results suggest that HCWs working in Hosp1 and Hosp2 face a higher risk when assigned to non-ICU, as compared to those assigned to the ICU; whereas HCWs in the ICU of Hosp3 face higher risk (see Fig. 1 and Table 3).

## DISCUSSION

Here, we studied the risk of incident SARS-CoV-2 infection among HCWs residing days and nights in quarantine hospitals, relying on detailed longitudinal data collected during the first wave of COVID-19 in Egypt. We found that most diagnosed infections (70%) occurred during what we defined as healthcare-associated outbreaks (as compared to isolated infections). We observed high variability in nosocomial incidence, ranging from 0.45 to 2.25 infections per 100 PD across the three hospitals. Using a model-based approach, we estimated the risk of both diagnosed and undiagnosed SARS-CoV-2 infections among HCWs, by hospital and by hospital unit (ICU and non-ICU). We further estimated that a substantial proportion of infections may have remained undetected, ranging from 45.0% to 59.2% across the three hospitals.

Our study design has several limitations. First, screening HCWs using serological tests before starting their working shifts may not detect active infections at the time of testing. Indeed, antibodies may be detected by serological tests in less than 40% of infected individuals within 7 days since symptoms onset, reaching higher detection levels at day 16 after symptom onset [17]. Hence, serological screening may allow recently infected HCWs –who are probably infectious– to start a working shift. However, the Egyptian public health authorities’ choice of a protocol relying on serological tests before working shifts was mainly driven by material constraints. Indeed, the use of rapid serological tests as a diagnostic tool was and remains of particular interest in limited-resource contexts due to their lower cost, the minimal equipment required, and faster results, as compared with RT-PCR tests [18]. Second, the assumption of a constant overall risk of infection among HCWs disregards dynamic fluctuations in risk driven by variation in the number of HCWs and/or patients in the hospitals, as well as the frequency and nature of HCWs’ contacts during their working shifts. This simplification was made in order to estimate average transmission rate first; fine reproduction of transmission dynamics were left for future work. Third, despite the quarantine strategy being adopted nationally, the differences between the organizations of each of the three hospitals (working shifts lengths, quarantine-organization period) resulted in some difficulties in comparing the risk estimations across the three hospitals. Data from other hospitals that have adopted a quarantine strategy will be beneficial to better understand occupational risks and eventually study the performance of such a strategy in comparison to standard care settings.

We found a ∼5-fold higher observed risk for Hosp3 as compared with the risk found in Hosp1; whereas the risk of infection at Hosp2 was just slightly higher. The higher risk of SARS-CoV-2 infection we report for Hosp3 and the higher risk faced by HCWs assigned to the ICU of Hosp3 may be partly explained by the short period over which this hospital adopted the quarantine organization, which coincided with the country’s highest epidemic activity of the first wave [4]. This may have led to a high proportion of severe and thus highly contagious COVID-19 patients referred to quarantine hospitals together with a higher workload for HCWs. Contrarily, Hosp1 adopted the quarantine organization in mid-March 2020, and thus experienced several weeks of lower epidemic intensity that may have improved preparedness for intense COVID-19 activity and implementation of infection control measures for invasive procedures (e.g., intubation in the ICU).

Our model-based approach, accounting for false-negativity rates of testing, as well as the right-truncation of our data, allowed us to estimate the risk of both diagnosed and undiagnosed SARS-CoV-2 infection among HCWs, by hospital and by hospital care unit. We estimated that a rigorous quarantine organization with systematic testing of HCWs at the end of their working shifts captures only about half of all HCW infections (depending on the length of the working shift). Of note, the higher proportion of infections that remained undetected in Hosp3 (59.2%, versus 46.4% and 45.0% in Hosp1 and Hosp2, respectively) may be explained by the shorter duration of the working shifts (7 days), which may result in false-negative tests for HCWs infected a short time before the end of their shift. Moreover, these results suggest that, in the absence of the quarantine-hospital organization and, more specifically, in the absence of systematic testing at the end of quarantine working shifts, an even larger proportion of infections among HCWs may remain undetected, thus putting HCWs’ close contacts at risk of infection [19] and, consequently, putting themselves at risk for adverse psychological symptoms [20].

The model-based estimates we found for the SARS-CoV-2 infection risk in Hosp1 and Hosp2 are consistent with infection point-prevalence reported in earlier studies performed in non-quarantine Egyptian hospitals (varying from 4.2% to 14.3%) and with incidence estimates reported among front-line HCWs in the UK (13% infection rate after one month) [10,11,21]. For comparison, a summary of previous results obtained early in the Covid-19 pandemic and/or specifically in the Egyptian context is presented in Table 4. Moreover, our results suggest that HCWs assigned to non-ICU face a higher risk than those assigned to the ICU, in Hosp1 and Hosp2, which is in line with what was observed in previous studies addressing the occupational risk of SARS-CoV-2 infection for HCWs in non-quarantine settings [22]. Overall, our findings on Hosp1 and Hosp2 suggest that, providing sufficient preparedness, HCWs working in quarantine hospitals may not face a higher infection risk, and thus highlight the benefits of implementing a quarantine-hospital strategy.

**Table 4.** Our results in a context. Our estimation of the per-shift probability of infection in Hosp1–3, along with estimates of the point-prevalence in other Egyptian studies and the observed cumulative incidence rate among HCWs in an English study.

| Indicator | Estimate (%) | Context | Study |
| --- | --- | --- | --- |
| **Per-shift probability of infection (95% CrI)** | 12.8 (7.6–19.5) | Hosp1 (14-days shift) | Our results |
|  | 17.3 (7.5–30.7) | Hosp2 (14-days shift) |  |
|  | 48.2 (23.8–74.5) | Hosp3 (7-days shift) |  |
| **Point-prevalence** | 4.2 | Symptomatic and asymptomatic HCWs in 12 healthcare facilities (N=4040). | [23] |
| **Point-prevalence** | 14.3 | Asymptomatic HCWs in emergency departments of tertiary care facility (N=203). | [10] |
| **Incidence rate** | 13.0 | Front-line HCWs testing negative at enrollment. | [21] |

A notable strength of our study lies in the specific nature of the quarantine hospital set-up: because HCWs resided continuously in the hospitals over their entire working shifts, we were able to exclude risk of infection in the community and specifically quantify the nosocomial risk for HCWs. Conversely, results reported from non-quarantine hospitals worldwide are generally unable to distinguish between the nosocomial vs. community risk of infection. However, as most studies conducted in healthcare settings, we were unable to distinguish between patient-to-HCW and HCW-to-HCW routes of transmission. A previous study on several nosocomial Covid-19 healthcare-associated outbreaks in Germany reported that HCW-to-HCW transmission could represent an outsized risk as compared to the one due to infected patients [24]. Still, assessing the relative contribution between patient-to-HCW versus HCW-to-HCW SARS-Cov-2 transmission was left for future work. However, it is worth noting that during the outbreaks observed in our study, infections occurred in different care units, which may prime HCW-to-HCW transmission, rather than simultaneous independent events of patient-to-HCW transmission. In early stages of the pandemic, personal protective equipment (PPE) placed a focus on the risk induced by patients, but could have contributes to underestimate the risk of infection from –infected and undiagnosed– colleagues. This may be especially true for quarantine hospitals in which HCWs share resting and conviviality rooms for longer times than in standard care settings. Adapting the use of PPE and social distancing between colleagues while ensuring sufficient social interaction and support to maintain HCWs’ mental health thus constitutes a specific challenge for quarantine hospitals [20].

Preventing nosocomial SARS-CoV-2 infection remains an urgent need, especially in settings where preparedness does not match the risk faced by HCWs. The large variation in infection risk we found between hospitals suggests that HCWs from quarantine hospitals may face a high risk of infection, but that, with sufficient anticipation and infection control measures, this risk can be brought down to levels similar to those observed in standard COVID-19 care settings. A comprehensive assessment of quarantine hospital care models should also include their impact on HCWs’ mental health as well as the potential benefits of earlier infection diagnosis, which is likely to reduce further hospital, household and community transmission [20].

## Data Availability

Please contact the corresponding author for access to the data

## EMAE-MESuRS working group on nosocomial SARS-CoV-2 modeling

Audrey Duval^6,7,8^, Kenza Hamzi^1^, Niels Hendrickx^6^, Kévin Jean^1,2,5^, Sofía Jijón^1,2^, Ajmal Oodally^6,7,1^, Lulla Opatowski^6,7^, George Shirreff^6,7,8^, David RM Smith^6,7,1^, Cynthia Tamandjou^7^ and Laura Temime^1,2^

^6^ Institut Pasteur, Epidemiology and Modelling of Antibiotic Evasion (EMAE), Paris, France

^7^Université Paris-Saclay, UVSQ, Inserm, CESP, Anti-infective evasion and pharmacoepidemiology team, Montigny-Le-Bretonneux, France

^8^IAME, UMR 1137, Université Paris 13, Sorbonne Paris Cité, France

## Declaration of interest

The authors declare no conflicts of interest.

## Funding

This study was supported by INSERM-ANRS (France Recherche Nord and Sud Sida-HIV Hépatites, grant number ANRS-COV-19).

## Acknowledgments

The authors thank all the dedicated HCWs who contributed to this study.

## Notes

### Competing Interest Statement

The authors have declared no competing interest.

### Funding Statement

This study was supported by INSERM-ANRS (France Recherche Nord and Sud Sida-HIV Hepatites, grant number ANRS-COV-19.).

### Author Declarations

The study was approved by the Research Ethics Committee (REC) of the Central Directorate of Research and Health Development and Reviews at the Egyptian Ministry of Health and Population (Serial: 25-2020/16), and by REC for human subject research at the Faculty of Medicine, Helwan University (Serial: 50-2020).

### Summary of Updates

Minor grammar edits

