## Supplementary Figure 1 for "Estimating the risk of incident SARS-CoV-2 infection among healthcare workers in quarantine hospitals: the Egyptian example" (from another version of this preprint: version 2, 2022-02-07)

### Supplementary figures

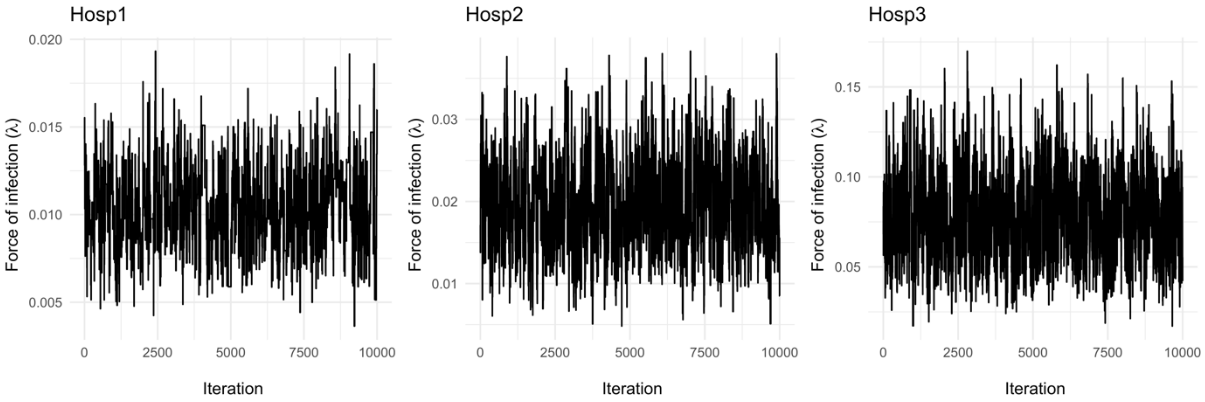


**Figure S1 Markov Chain Monte-Carlo diagnostics**
Trace plot of the posterior distribution for the force of infection ($\lambda$), for Hosp1–3. In 10,000 iterations, the sample space is explored many times and no high serial correlation is apparent in the chain.
